# COVID-19 vaccine uptake and acceptability during the second phase of vaccine rollout: a community-based survey among household heads in Zamfara state, Nigeria, 2021

**DOI:** 10.1101/2023.12.14.23299963

**Authors:** Ahmad Suleiman Idris, Hafiz Aliyu, Rabi Usman, Ahmed Tijani Abubakar, Muhammad Abdullahi Maijawa, Bello Suleiman Abdullahi, Muhammad Shakir Balogun, Abdulhakeem Olorukooba, Chukwuma Umeokonkwo, Abubakar Maiyaki, Muhammad Sani, Muhammad Yisa, Ismail Hadi Zubair, Onu Hastings Chinedu, Tenmuso David Jatau, Kabir Sabitu

## Abstract

**Introduction:** Vaccines have played an important role in the control of infectious diseases globally. COVID-19 vaccine was rolled out in Zamfara State for the public in August 2021. We aimed to determine the level of COVID-19 vaccine uptake, acceptance, and awareness in Zamfara state Nigeria, during the second phase of the COVID-19 roll-out in the community.

**Methods:** We executed a descriptive cross-sectional study in Zamfara state, Nigeria. We used multistage sampling technique to randomly select 910 household heads between October 12 to December 20, 2021. We used a semi-structured electronic questionnaire to collect data on sociodemographic characteristics, uptake, and acceptance of COVID-19 vaccine. We performed descriptive analysis; calculated frequencies, proportions, and produced tables and figures.

**Results:** Our 899 respondents had a median age of 48 years (interquartile range: 29.5-66.5). About 78.1% (711) were males. A majority of the respondents were within the age group 50-59 years. Of 897 respondents 47.2% (423) were educated up to the secondary school level.

Only 8.9% (81) had received COVID-19 vaccine. Of the 829 unvaccinated respondents, 10.1% (84) accepted to take the vaccine the current week of the interview, 38.4% (318) would it the following week, and 27.4% (227) the following month, while 12.2% (101) of the respondents rejected the vaccine,

**Conclusion:** COVID-19 vaccine had a poor uptake and acceptance rate in Zamfara State during the vaccine rollout. We recommended carefully designed and targeted sensitization campaigns to increase the demand of COVID-19 vaccine in the community.

## Introduction

### Background

The world recorded a drop in vaccine uptake since the COVID-19 pandemic began.^[10]^ It is paramount to investigate COVID-19 vaccine uptake in Nigeria when there is drop in most countries.^[10]^ Peculiar factors distinguish this infectious disease from other re-emerging diseases.^[10]^

Over 158 candidates of COVID-19 vaccine were developed through the Access to COVID-19 Tools (ACT) Accelerator which is a global collaboration to accelerate development, production, and equitable access to COVID-19 tests, treatments, and vaccines.^[12]^ The first, vaccines pillar led by CEPI, Gavi (the Vaccine Alliance) and the WHO, to facilitate collaboration and increase the speed of the search for an effective COVID-19 vaccine and ensure its’ delivery globally. At the same time, supporting the building of manufacturing capabilities and buying supply, ahead of time, so that COVID-19 vaccine doses can be distributed fairly in the places of greatest need, worldwide. Hence, vaccination is our best chance towards ending the pandemic.

Previous studies on vaccine acceptance and theories of health behaviour, such as the health belief model or protection motivation theory, have identified many factors that influence the acceptance or uptake of a pandemic vaccine, including the risk perception of the disease, perception of vaccine safety and efficacy, general vaccination attitude, past vaccination history, recommendations from doctors, price, vaccination convenience and socio-demographic characteristics.^[13]^

In a recent study conducted in Cameroon, the researchers found that more than 80% of the sample population had good knowledge of COVID-19, 69% had a positive attitude, and 60% adopted preventive practices. In north-central Nigeria, a cross-sectional study found that 99.5% of respondents had good knowledge of COVID-19, about 80% had positive attitudes towards adhering to government Infection Prevention and Control (IPC) measures, and more than 90% adopted preventive behaviours like social distancing, improved personal hygiene, and using face mask. The willingness of people to adopt preventative public health behaviours is well known to influence spread of disease, and these are often associated with public risk perception.^[14]^

In a large study conducted to assess the public risk perception of COVID-19 around the world, national samples were obtained from countries across Europe, America, and Asia.^[14]^ The researchers found that several factors were significant predictors of risk perception in more than half of the countries examined. ^[14]^These include, personal experience with the virus, social amplification by family and friends, prosocial values, and individualistic worldviews.^[14]^ Trust in government, and science and medical professionals were also significant predictors of risk perception.^[14]^ In another study conducted among college students in China, researchers found a high risk perception, which was significantly influenced by gender, location (schools located in Hubei), and knowledge level.^[15]^

Vaccine acceptability has been a problem since the discovery of vaccines and that of COVID-19 is not likely to be an exception.^[16]^ In an online study done in South-East Asia on a hypothetical COVID-19 vaccine acceptability revealed an acceptance of 93.3% when perceived efficacy of 95% was used, but only to drop to 67.0% when the vaccine efficacy of 50% as used.^[17]^ Even though this study was conducted in a population with a literacy level above 90%, excluding populations like farmers that rarely use the internet, a significant drop was still recorded when perception of efficacy dropped.^[17]^ Willingness to accept a vaccine ranged from about 90% in China to less than 60% in Russia; with an average of 71.5% if the vaccine was perceived to be safe and efficacious.^[18]^ Factors influencing willingness to accept vaccine include trust in the central government, and employer’s recommendation.^[18]^ Other factors include age, gender, income, and level of education, although these factors didn’t show a strong association.^[18]^ Evidence of willingness to vaccinate against coronavirus disease (COVID-19) is insufficient in 19 countries that were surveyed, highlighting the need for targeted interventions to increase and sustain public acceptance of an incoming COVID-19 vaccine.^[18]^ Our aim was to determine the factors associated with the awareness, uptake, and acceptability of COVID-19 vaccine among the general population of Gusau metropolis, Zamfara State.

## Methods

### Study Area

The study area is Gusau, the capital of Zamfara State in north-western Nigeria. Zamfara state has 14 local government areas (LGAs) within a land area of 37,931 Km2. Zamfara has a population of about 5,307,154 and the people are mainly farmers.

### Study Design

This was a cross-sectional study.

### Study Population

The study participants were drawn from household heads residing in Gusau and Bungudu LGA of Zamfara State. While those living in security compromised areas were be excluded from the study.

### Sample Size determination

The sample size was calculated for the state with the household as the sampling frame. At 95% confidence level, 5% margin of error, population proportion of 80.0% (0.8) from a previous Nigerian vaccine acceptance study ^[16]^, and an unlimited population size, *Z* is 1.96

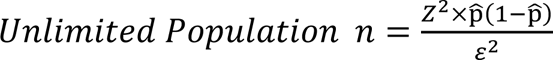

Where,

***z* is** the *z* score

**ε** is the margin of error

**n** is sample size

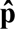 is the population proportion

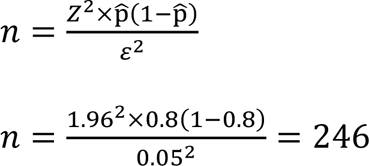

Using non-response rate of 7% in a similar Nigerian study^[16]^

= 246 + (0.07 × 246)

= 264 people

### Sampling Technique

A multistage sampling was used. In the first stage, 9 wards were selected in Gusau and Bungudu LGA after excluding security compromised wards. In the second stage, 10 settlements were randomly selected from each ward. In the third stage, the first household in each settlement was randomly selected and the remaining households were selected by skipping 10 households. The heads of the household were interviewed after getting consent, 11 households were selected in each settlement, and a total of 990 households were selected for the study.

### Data Collection Tools

A standardised semi-structured electronic questionnaire was used to collect data on the sociodemographic characteristics, knowledge on COVID 19, uptake, awareness, and acceptability of COVID-19 vaccine.

### Data Collection Procedure

Data was collected between October 12 to December 20, 2021. Data collectors were trained to administer the questionnaire in both Hausa and English. The questionnaire was pretested in a ward that was not be included in the main study. The modified pretested questionnaire was used to collect the final data from the household heads. The respondents with poor knowledge or no awareness of COVID-19 or its vaccine were sensitised by the data collectors before completion of the survey.

### Data Analysis

Data was analysed using Stata 17BE and Microsoft Excel. After cleaning and coding, descriptive analysis was done to determine the frequencies of the sociodemographic characteristics like the age, sex, place of residence, educational status respondents, testing status for COVID-19 before the study. Also, data on the knowledge of COVID-19, COVID-19 vaccination status, acceptability of COVID-19 vaccine, uptake of routine immunization vaccines, and trust of the respondents was analysed.

### Variable Measurement

The attitude of the respondents towards was measured using a 5 point Likert scale with the options strongly agree, agree, neutral, disagree, and strongly disagree. Also, the level of trust was measured with a 5 point Likert scale with options very much, much, some, little, and very little.

### Ethical Considerations

An ethical clearance for the study was obtained from Zamfara State Ministry of Health before commencement. Only participants that gave consent were part of the study. No identifying information was collected from the participants. All participants lacking in knowledge of COVID-19 or its vaccine were sensitised before conclusion of the study. Data from the study was secured in a password protected computer and only authorised personnel have access to it.

## Results

This is a pilot study that provided the most current information on the level and factors associated with acceptability of COVID-19 vaccine in Gusau metropolis, Zamfara State during the vaccine rollout in September 2021. We approached 990 respondents, 91.9% (910) of them gave consent and responded to our questions, and we had a non-response rate of 8.1%. Of the 910 respondents, 11 had missing or invalid ages and were excluded in the analysis for age.

### Descriptive Statistics

#### Sociodemographic characteristics

The respondents had a median age of 48 years with an interquartile range of 18. Males made up 78.1% (711) of the respondents. Majority of the respondents were within the 50-59 years category. Of 897 respondents 47.2% (423) were educated up to the secondary school level, 7.4% (66) received no education, 4.5% (40) received an undergraduate degree, and 0.3% (3) received a graduate or professional degree. The Hausa and Muslims made up 97.9% (891) each of the respondents. About 95.5% (869) of the respondents were married. Also, 89.2% (812) of the respondents were employed with the majority 44.6% (362) of them being farmers with healthcare workers making about 4.2% (34) of the employed respondents. Among 910 of the respondents, 11.2% (102) of them had at least one chronic illness. Table 1 below shows the details of the sociodemographic characteristics of the respondents.

**Table 1:**
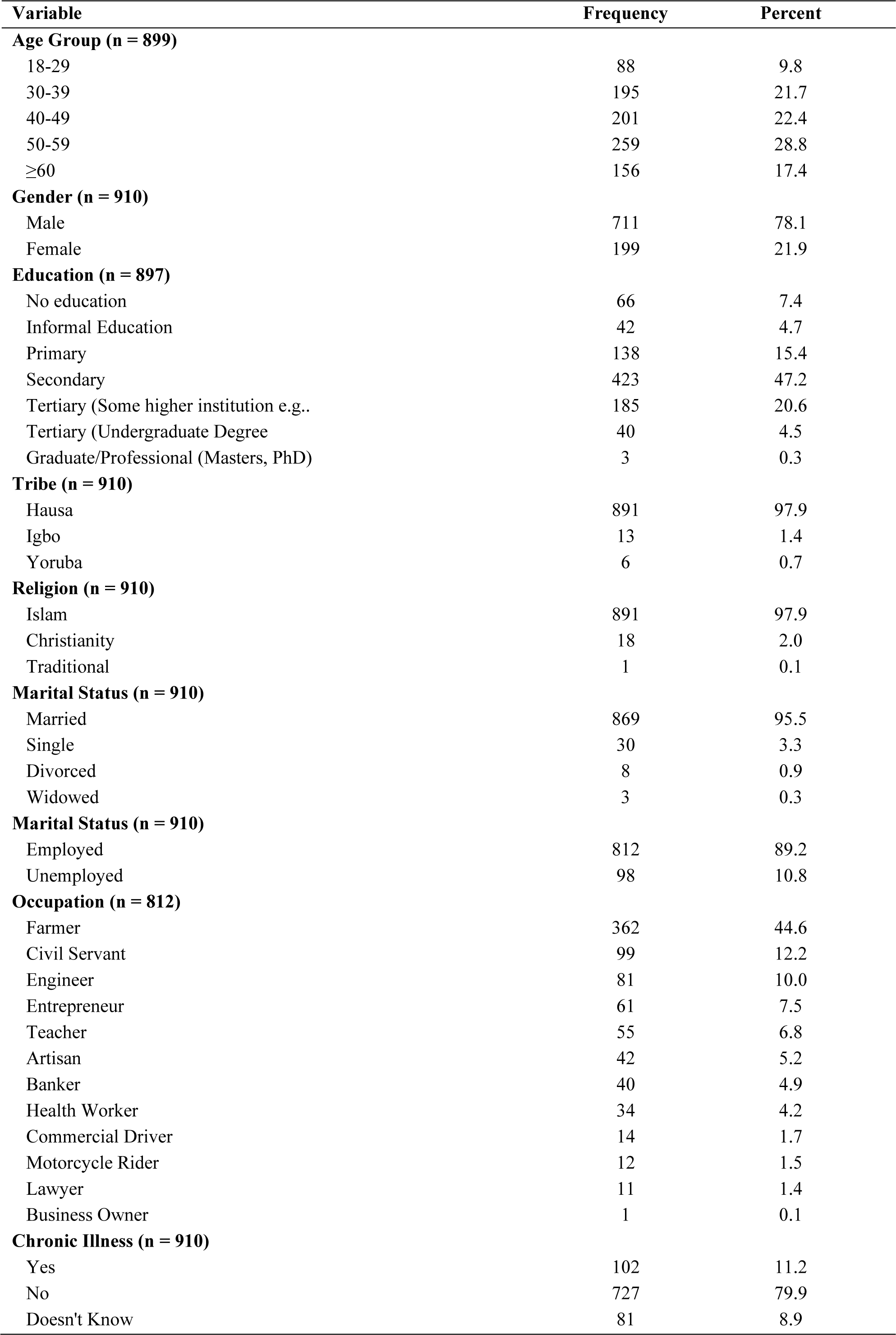
Sociodemographic characteristics of the respondents.

| Variable | Frequency | Percent |
| --- | --- | --- |
| **Age Group (n = 899)** |  |  |
| 18-29 | 88 | 9.8 |
| 30-39 | 195 | 21.7 |
| 40-49 | 201 | 22.4 |
| 50-59 | 259 | 28.8 |
| ≥60 | 156 | 17.4 |
| **Gender (n = 910)** |  |  |
| Male | 711 | 78.1 |
| Female | 199 | 21.9 |
| **Education (n = 897)** |  |  |
| No education | 66 | 7.4 |
| Informal Education | 42 | 4.7 |
| Primary | 138 | 15.4 |
| Secondary | 423 | 47.2 |
| Tertiary (Some higher institution e.g.. | 185 | 20.6 |
| Tertiary (Undergraduate Degree | 40 | 4.5 |
| Graduate/Professional (Masters, PhD) | 3 | 0.3 |
| **Tribe (n = 910)** |  |  |
| Hausa | 891 | 97.9 |
| Igbo | 13 | 1.4 |
| Yoruba | 6 | 0.7 |
| **Religion (n = 910)** |  |  |
| Islam | 891 | 97.9 |
| Christianity | 18 | 2.0 |
| Traditional | 1 | 0.1 |
| **Marital Status (n = 910)** |  |  |
| Married | 869 | 95.5 |
| Single | 30 | 3.3 |
| Divorced | 8 | 0.9 |
| Widowed | 3 | 0.3 |
| **Marital Status (n = 910)** |  |  |
| Employed | 812 | 89.2 |
| Unemployed | 98 | 10.8 |
| **Occupation (n = 812)** |  |  |
| Farmer | 362 | 44.6 |
| Civil Servant | 99 | 12.2 |
| Engineer | 81 | 10.0 |
| Entrepreneur | 61 | 7.5 |
| Teacher | 55 | 6.8 |
| Artisan | 42 | 5.2 |
| Banker | 40 | 4.9 |
| Health Worker | 34 | 4.2 |
| Commercial Driver | 14 | 1.7 |
| Motorcycle Rider | 12 | 1.5 |
| Lawyer | 11 | 1.4 |
| Business Owner | 1 | 0.1 |
| **Chronic Illness (n = 910)** |  |  |
| Yes | 102 | 11.2 |
| No | 727 | 79.9 |
| Doesn't Know | 81 | 8.9 |

#### Uptake and acceptance of COVID-19 vaccine, previous history of any vaccination

Table 2 below shows the COVID-19 vaccine uptake and level of acceptance among those that had not received the vaccine yet.

**Table 2:** Vaccine uptake and acceptance among the respondents.

| **Variable** | **Frequency** | **Percent** |
| --- | --- | --- |
| **COVID-19 Vaccination Status (n = 910)** |  |  |
| Vaccinated | 81 | 8.9 |
| Unvaccinated | 829 | 91.1 |
| **Reasons for taking COVID-19 Vaccine** |  |  |
| It was recommended by my colleagues | 21 | 25.9 |
| It was recommended by my doctor | 24 | 29.6 |
| I took it on my own to protect myself | 19 | 23.5 |
| My boss demanded I should take it | 18 | 22.2 |
| Vaccine saves life | 13 | 16.1 |
| No reason | 3 | 3.7 |
| **Reasons for not accepting the COVID-19 vaccine** |  |  |
| The vaccine will not protect me | 34 | 33.7 |
| The vaccine was made too fast | 21 | 20.8 |
| The vaccine is not safe | 13 | 12.9 |
| The vaccine contains microchips | 10 | 9.9 |
| I want some people to take it first to see if nothing bad happens | 41 | 40.5 |
| It is prohibited by my religion | 1 | 2.0 |
| No reason | 4 | 4.0 |
| **Acceptance of COVID-19 vaccine** |  |  |
| I don't know | 46 | 5.6 |
| No | 101 | 12.2 |
| Yes, next month | 227 | 27.4 |
| Yes, next week | 318 | 38.4 |
| Yes, next year | 53 | 6.4 |
| Yes, this week | 84 | 10.1 |
| **Ever received any vaccine in the past? (n = 910)** |  |  |
| No | 67 | 7.4 |
| Yes | 770 | 84.6 |
| Don't know | 73 | 8.0 |
| **Reasons for not ever taking any vaccine in the past (n = 67)** |  |  |
| I don't believe in Immunization | 23 | 34.3 |
| I don't know the benefit | 11 | 16.4 |
| I don't know where to get it | 14 | 20.9 |
| I don't know the benefit | 11 | 16.4 |
| I don't like the attitude of HCWs | 10 | 14.9 |
| I was not taken for immunization as a child | 13 | 19.4 |
| My Religion does not allow it | 11 | 16.4 |

#### Uptake

About 8.9% (81) of the respondents had received the COVID-19 vaccine. Among those that took the vaccine, the reasons for uptake for 29.6% (24) was recommendation by doctor, 25.9% (21) by colleagues, 23.5% (19) took it on their own for protection, 22.2% (18) because their boss demanded, while 16.1% (13) believed it saves lives, and 3.7% (3) had no reason for taking the vaccine.

#### Acceptance

Of the 829 unvaccinated respondents, 10.1% (84) accepted taking COVID-19 vaccine the current week of the interview, 38.4% (318) the following week, 7.4% (227) the following month, 6.4% (53) the following year, while 12.2% (101) would not take the vaccine, and 5.6% (46) don’t know what they would do. The 101 respondents that confirmed they would not take the vaccine were asked for the reasons why they made the response. For the majority, 40.5% (41) want some people to take it first to observe its safety, 3.7% (34) believe the vaccine will not protect them, 20.8% (21) think the vaccine was made too fast, 12.9% (13) believe the vaccine is not safe, 9.9% (10) believe the vaccine contains microchips, while 4% (4) had no reason.

#### Awareness of COVID-19 pandemic and availability of COVID-19 vaccine

The respondents were asked about their awareness of COVID-19 Pandemic and COVID-19 vaccine availability; 95.8% (872) and 84.2% (766) were aware of the pandemic and the COVID-19 vaccine availability respectively. The 766 respondents aware of the COVID-19 vaccine were asked about their sources of information and 87.7% (672) found out through the radio, 35.95% (275) through the social media, 14.2% (109) through the television, 12.1% (93) through family and friends, 4.1% (31) through websites, 2.2% (17) through healthcare professionals, and 0.5% (4) through health officials (NCDC).

**Figure 1:**
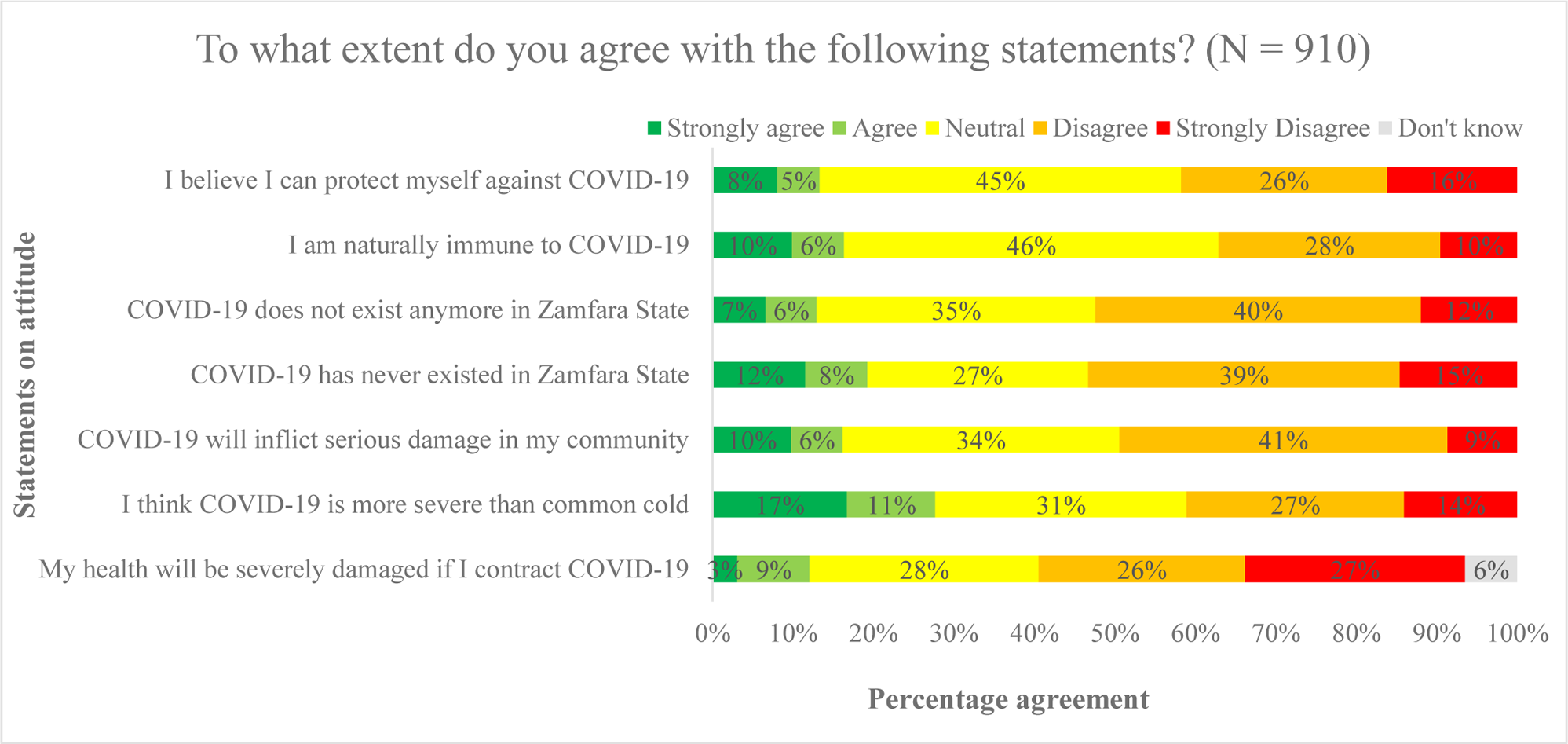
The attitude of the population towards the COVID-19 pandemic.

**Table 3:** Awareness of COVID-19 pandemic and COVID-19 vaccine in the respondents.

| Variable | Frequency | Percent |
| --- | --- | --- |
| **Awareness COVID-19 Pandemic (n = 910)** |  |  |
| Yes | 872 | 95.8 |
| No | 38 | 4.2 |
| **How did you get to know about the vaccine? (n = 766)** |  |  |
| Radio | 672 | 87.7 |
| Social Media | 275 | 35.9 |
| Television | 109 | 14.2 |
| Family/Friends | 93 | 12.1 |
| Newspapers and Magazines | 31 | 4.1 |
| Websites | 21 | 2.7 |
| Healthcare Professionals | 17 | 2.2 |
| Healthcare Officials (NCDC) | 4 | 0.5 |
| **Ever Tested for COVID-19 (n = 872)** |  |  |
| No | 567 | 65.0 |
| Yes, don't know my result yet | 111 | 12.7 |
| Yes, negative result | 106 | 12.1 |
| Yes, Positive | 88 | 10.0 |
| **Awareness COVID-19 Vaccine Availability (n = 910)** |  |  |
| Yes | 766 | 84.2 |
| No | 144 | 15.8 |

#### Attitude, trust, and confidence

As shown in table 4 and figure 2, most respondents 28.5% (259) were either “neutral” or “disagreed” 25.6% (233) with the statement *my health will be severely damaged if I contract COVID-19.* Also, 31.2% (284) were either “neutral” or “disagreed” 27% (246) with the statement *I think COVID-19 is more severe than common cold*. Similarly, 34.4% (313) were either “neutral” or “disagreed” 40.8% (371) with the statement *COVID-19 will inflict serious damage in my community.* Also, 27.5% (250) were either “neutral” or “disagreed” 38.7% (352) with the statement *COVID-19 has never existed in Zamfara State.* Also, 34.6% (315) were either “neutral” or “disagreed” 40.4% (368) with the statement *COVID-19 does not exist anymore in Zamfara State*.

**Figure 2:**
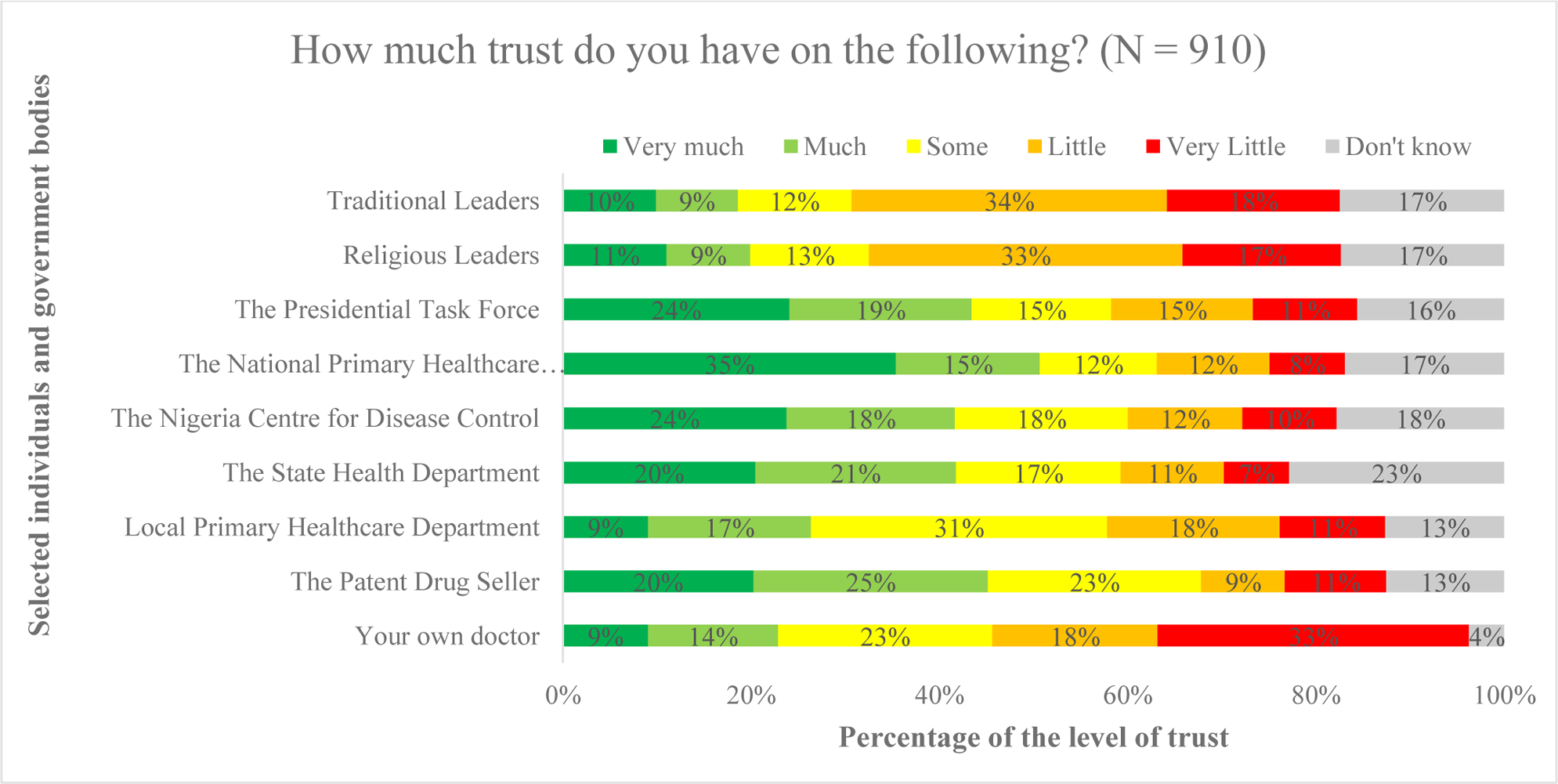
The level of trust the population has on individuals and some government bodies.

**Table 4:** Attitude of the respondents towards COVID-19 and COVID-19 vaccine.

| Question (n=910) | Strongly Agree (%) | Agree (%) | Neutral (%) | Disagree (%) | Strongly Disagree (%) | Don't Know (%) |
| --- | --- | --- | --- | --- | --- | --- |
| **To what extent do you agree or disagree on the following statements?** |  |  |  |  |  |  |
| My health will be severely damaged if I contract COVID-19 | 28 (3.1) | 82 (9.0) | 259 (28.5) | 233 (25.6) | 249 (27.4) | 59 (6.5) |
| I think COVID-19 is more severe than common cold | 152 (16.7) | 100 (11.0) | 284 (31.2) | 246 (27.0) | 128 (14.1) | 0 (0.0) |
| COVID-19 will inflict serious damage in my community | 89 (9.8) | 58 (6.4) | 313 (34.4) | 371 (40.8) | 79 (8.7) | 0 (0.0) |
| COVID-19 has never existed in Zamfara State | 105 (11.5) | 70 (7.7) | 250 (27.5) | 352 (38.7) | 133 (14.6) | 0 (0.0) |
| COVID-19 does not exist anymore in Zamfara State | 60 (6.6) | 58 (6.4) | 315 (34.6) | 368 (40.4) | 109 (12.0) | 0 (0.0) |
| I am naturally immune to COVID-19 | 90 (9.9) | 59 (6.5) | 423 (46.5) | 251 (27.6) | 87 (9.6) | 0 (0.0) |
| I believe I can protect myself against COVID-19 | 73 (8.0) | 48 (5.3) | 409 (45.0) | 233 (25.6) | 147 (16.2) | 0 (0.0) |

In table 5, majority 33.1% (301) and 22.8% (207) of the respondents had “Very Little” and “Some” trust on their doctors respectively. And 25% (227) and 22.7% (206) had “Much” and “Some” trust in the patent drug seller respectively. Also, 31.4% (286) and 18.4% (167) had “Some” and “Little” trust in the local primary healthcare department. While 21.3% (194) and 20.4% (186) had “Much” and “Very Much” trust in the state health department respectively. The respondents had 23.7% (216) and 18.4% (167) had “Very Much” and “Some” trust in the Nigeria Centre for Disease Control respectively. Most of the respondents 35.4% (322) had “Very Much” trust in the National Primary Healthcare Development Agency, 24.1% (219) had “Very Much” trust the Presidential Task Force. While 33.3% (303) and 33.5% (305) had “Little” trust in both Religious and Traditional leaders respectively. When asked to rank the above individuals and government bodies. The first choice to the eight choice, first being the best. The respondents rated their doctor (38%), patent drug seller (29%), the state health department (38%), Nigeria Centre for Disease Control (31%), National Primary Healthcare Development Agency (30%), Religious Leaders (55%), and Traditional Leaders (57%) as the first to eight choices respectively. The details of the ranking are shown in figure 3.

**Figure 3:**
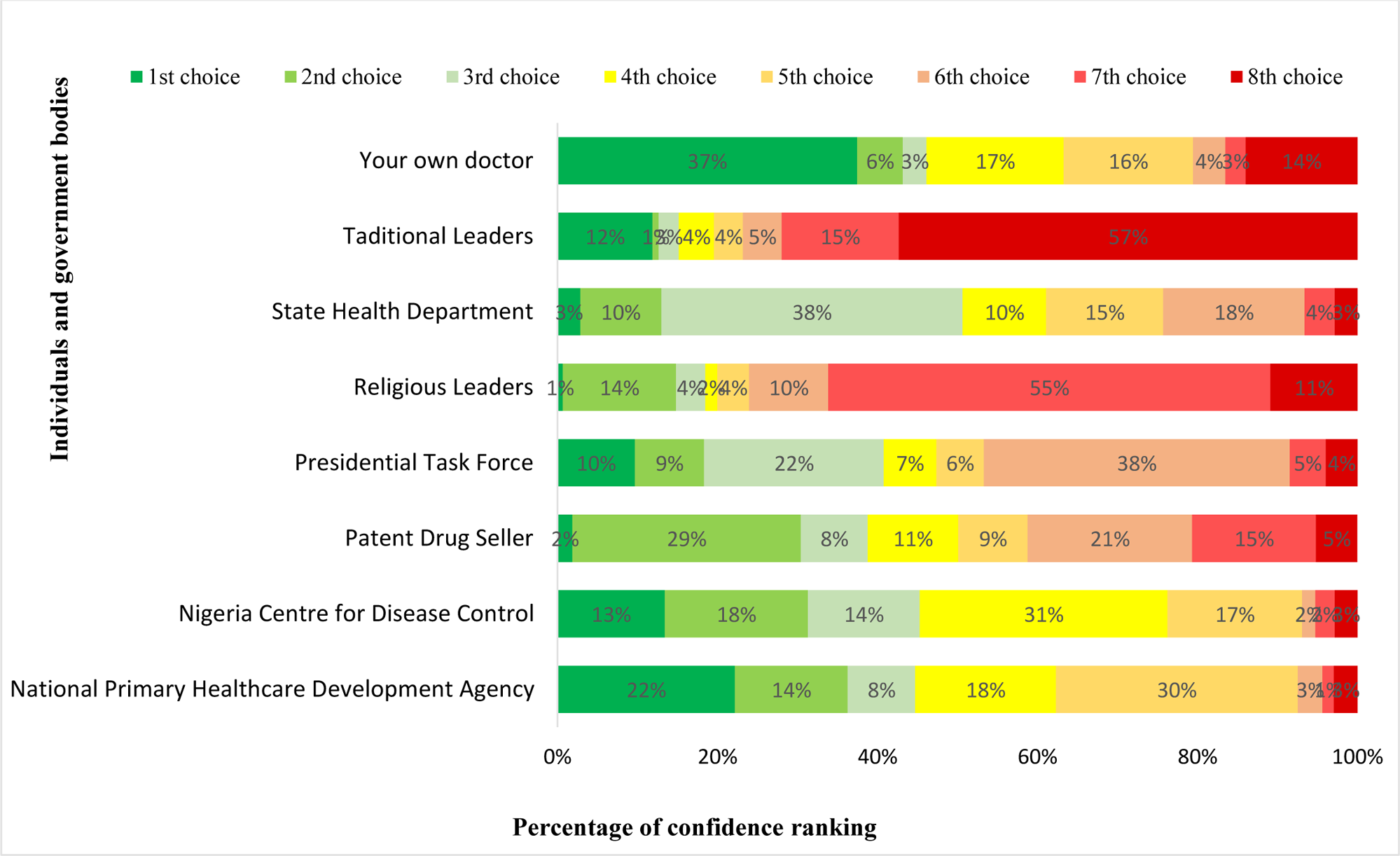
The level of confidence in individuals and government bodies (N = 910)

**Table 5:** The level of trust in individuals, leaders, and health institutions among the respondents.

| Question (n=910) | Very Much (%) | Much (%) | Some (%) | Little (%) | Very Little (%) | Don't know (%) |
| --- | --- | --- | --- | --- | --- | --- |
| **How much trust do you have on the following?** |  |  |  |  |  |  |
| Your own doctor | 82 (9.0) | 126 (13.9) | 207 (22.8) | 160 (17.6) | 301 (33.1) | 34 (3.7) |
| The Patent Drug Seller | 184 (20.2) | 227 (25.0) | 206 (22.6) | 81 (8.9) | 98 (10.8) | 114 (12.5) |
| Local Primary Healthcare Department | 82 (9.01) | 158 (17.4) | 286 (31.4) | 167 (18.4) | 102 (11.2) | 115 (12.6) |
| The State Health Department | 186 (20.44) | 194 (21.3) | 159 (17.5) | 100 (11.0) | 63 (6.9) | 208 (22.9) |
| The Nigeria Centre for Disease Control | 216 (23.74) | 163 (17.9) | 167 (18.4) | 111 (12.2) | 91 (10.0) | 162 (17.8) |
| The National Primary Healthcare Development Agency | 322 (35.39) | 139 (15.3) | 113 (12.4) | 109 (12.0) | 73 (8.0) | 154 (16.9) |
| The Presidential Task Force | 219 (24.07) | 176 (19.3) | 135 (14.8) | 137 (15.1) | 101 (11.1) | 142 (15.6) |
| Religious Leaders | 100 (10.99) | 81 (8.9) | 115 (12.6) | 303 (33.3) | 153 (16.8) | 158 (17.4) |
| Traditional Leaders | 90 (9.90) | 79 (8.7) | 110 (12.1) | 305 (33.5) | 167 (18.4) | 159 (17.5) |

## Discussion

This study presents the latest findings from a community-based survey rather than the regular online surveys that have been published from Nigeria. Many factors were considered in the design and execution of the study. Our study highlights the uptake of COVID-19 vaccine, and it’s acceptability by the general population of Gusau metropolis in Zamfara State. It also highlighted some key issues in awareness of COVID-19, its vaccine, and attitudes of the general population towards COVID-19 and its vaccine.

In our study, most of the respondents were males and 50 years and older. This was from the design because we interviewed the heads of the house households given the setup of a typical Northern Hausa community and the extended family system, the eldest male in most cases, is the head of the household.^[19]^ Almost half of our respondents have their highest level of education at the secondary school level. This is the situation in most Northern states of Nigeria where education is generally poor.^[20]^ The Hausa-Muslims made up most of our respondents as most of the population in Zamfara State are Hausa-Muslims. Also, almost half of our respondents were farmers, as most of the population of Zamfara are farmers.^[21]^ About one-eight of out respondents have a chronic illness. This may be explained by the age of the respondents being more above fifty years as chronic illnesses tend to manifest more with increasing age.^[22]^

The uptake of COVID-19 in our study was low, our study was done during the peak of the roll out of the first dose of COVID-19 vaccine in Nigeria. It is like some studies conducted in the United States of America, Kuwait, Jordan, Italy, Russia, Poland, and France where the uptake was low.^[23,24]^ But this is in contrast with other studies in Ecuador, Malaysia, Indonesia, and China that reported a higher uptake above 90%.^[24]^ Among the reasons for uptake include recommendation by a doctor, self-motivated for protection, boss at work demanding uptake, strong belief vaccine saves life, and no reason at all for a very few. These reasons are similar to motivations for uptake as discussed by other studies.^[25,26]^

Studies on acceptance of COVID-19 vaccine have been conflicting due to many reasons. Population, method of questionnaire administration, knowledge, and even country. Most studies used a yes or no question to assess acceptance.^[23,24,26]^ Our study considered different levels of hesitancy. Any person that heard about COVID-19 vaccine availability and had access to it and didn’t take has some element of hesitancy.^[27–30]^ We measured different levels of hesitancy. Those that responded they would take the vaccine within the week were classified as non-hesitant. Those that would take the vaccine the following week were mildly hesitant, those that responded they won’t take it till next month were moderately hesitant, those that said they would take it next year were severely hesitant. Then we had the ones that were never going to take the vaccine, total rejection, and those that didn’t know were classified as unsure.

The strength of this study lies in its design, use of random sampling, and a large sample size. A pretested electronic questionnaire was used to minimize errors in data collection. Data collectors were trained to minimize bias. This study is the first of its kind in Zamfara State, it is the most current and probably the only baseline study on COVID-19 vaccine acceptability and uptake. As a pilot study, it will inform future similar studies in Zamfara State and Nigeria.

Our major limitations in this study are the fact that this is a cross-sectional study. Secondly, the study had a major drawback, during data collection, communication was lost between the researchers and data collectors because network was shut down by the government for 3 months due to frequent violent attacks by bandits in the state.

## Conclusion

COVID-19 vaccine had a poor uptake and acceptance rate in Zamfara State during the vaccine rollout in the community. The study findings also show various levels of COVID-19 vaccine hesitancy in the community. We recommended carefully designed and targeted sensitization campaigns to increase the demand of COVID-19 vaccine in the community.

## Data Availability

All data produced in the present study are available upon reasonable request to the authors

## Notes

### Competing Interest Statement

The authors have declared no competing interest.

### Funding Statement

This study did not receive any funding

### Author Declarations

Ethical approval was granted by the Zamfara State Ministry of Health.

